# Proteomic profiling of bronchoalveolar lavage fluid uncovers protein clusters linked to survival in idiopathic forms of interstitial lung disease

**DOI:** 10.1101/2024.05.30.24308215

**Authors:** Linh T. Ngo, Michaella J. Rekowski, Devin C. Koestler, Takafumi Yorozuya, Atsushi Saito, Imaan Azeem, Alexis Harrison, M. Kristen Demoruelle, Jonathan Boomer, Bryant R. England, Paul Wolters, Philip L. Molyneaux, Mario Castro, Joyce S. Lee, Joshua J. Solomon, Koji Koronuma, Michael P. Washburn, Scott M. Matson

**Author notes:** Corresponding author: Scott M. Matson, Assistant Professor of Medicine University of Kansas School of Medicine 4000 Cambridge St. Mailstop 3007. Authors contributed equally (Linh T. Ngo, Michaella J. Rekowski).

## Abstract

**Background:** Idiopathic interstitial pneumonias (IIPs) such as idiopathic pulmonary fibrosis (IPF) and interstitial pneumonia with autoimmune features (IPAF), present diagnostic and therapeutic challenges due to their heterogeneous nature. This study aimed to identify intrinsic molecular signatures within the lung microenvironment of these IIPs through proteomic analysis of bronchoalveolar lavage fluid (BALF).

**Methods:** Patients with IIP (n=23) underwent comprehensive clinical evaluation including pre-treatment bronchoscopy and were compared to controls without lung disease (n=5). Proteomic profiling of BALF was conducted using label-free quantitative methods. Unsupervised cluster analyses identified protein expression profiles which were then analyzed to predict survival outcomes and investigate associated pathways.

**Results:** Proteomic profiling successfully differentiated IIP from controls. *k*-means clustering, based on protein expression revealed three distinct IIP clusters, which were not associated with age, smoking history, or baseline pulmonary function. These clusters had unique survival trajectories and provided more accurate survival predictions than the Gender Age Physiology (GAP) index (C-index 0.794 vs. 0.709). The cluster with the worst prognosis featured decreased inflammatory signaling and complement activation, with pathway analysis highlighting altered immune response pathways related to immunoglobulin production and B cell-mediated immunity.

**Conclusions:** The unsupervised clustering of BALF proteomics provided a novel stratification of IIP patients, with potential implications for prognostic and therapeutic targeting. The identified molecular phenotypes underscore the diversity within the IIP classification and the potential importance of personalized treatments for these conditions. Future validation in larger, multi-ethnic cohorts is essential to confirm these findings and to explore their utility in clinical decision-making for patients with IIP.

## Background

Idiopathic interstitial pneumonia (IIP) encompasses a group of lung diseases with unknown causes that are distinct from those associated with autoimmune diseases or known antigen exposures. Among these, idiopathic pulmonary fibrosis (IPF) is the most common and is diagnosed using a multidisciplinary approach that integrates clinical, radiographic, and histopathologic criteria [1]. However, there exists a subset of IIP patients whose features diverge from the established IPF criteria, particularly those exhibiting autoimmune characteristics without meeting the criteria for a definitive autoimmune diagnosis, drawing significant interest in clinical and research settings. An international task force in 2015 defined this unique class of IIP as interstitial pneumonia with autoimmune features (IPAF) [2]. Patients with IPAF exhibit at least two autoimmune features from defined clinical, serological, and morphological domains [2]. Studies have suggested that IPAF patients may have a survival advantage over those with IPF, supporting the theory that IPAF represents a distinct clinical entity [3]. Given the presumed importance of immune dysregulation in IPAF and the uncertain balance of harms and benefits between immunomodulation and antifibrotic therapy for patients with IIP, there is significant interest in determining optimal treatment approaches for these patients [4].

In the absence of predictive markers for treatment response in patients with IPAF, clinicians are forced to extrapolate therapeutic decisions from anecdotal experiences with similar patients and from conditions with similar radiographic or clinical features. For instance, patients with IPAF whose radiographic pattern is predominantly fibrotic such as usual interstitial pneumonia (UIP) [5] or pleuroparenchymal fibroelastosis (PPFE) [6, 7] are more likely to be managed like patients with idiopathic pulmonary fibrosis, while IPAF patients with inflammatory radiographic patterns that resemble autoimmune interstitial lung disease (ILD) such as organizing pneumonia (OP) or non-specific interstitial pneumonia (NSIP) [8, 9] are often managed like those with established connective tissue-related interstitial lung disease (CTD-ILD). Given the importance of treatment considerations for these patients, it is imperative to better define the distinction across these idiopathic conditions based on intrinsic and meaningful molecular signatures.

Given the diversity of patients within the IIP classification schema, we hypothesized that lung microenvironment protein expression profiles would identify patient-specific clusters that influence clinical outcomes. We tested this hypothesis with an agnostic approach: by performing unsupervised clustering of IIP patients’ protein expression profiles followed by an examination of the survival trajectories of these clusters.

## Methods

### IIP Cohort

Patients with IPF and IPAF were collected at a single center between June 2013 and May 2017 for a prospective research protocol at the Department of Respiratory Medicine and Allergology at the Sapporo Medical University Hospital, Japan [10]. Utilizing published European Respiratory Society/American Thoracic Society IPAF research criteria [2], patients were identified from the parent registry by two board-certified pulmonologists who specialize in ILD. Patients were also evaluated upon enrollment in the parent registry by a rheumatologist who excluded the diagnosis of systemic autoimmune disease. Patients with IPF were identified from the same registry based on the most recently published international guidelines [1] via the same team of researchers. The Institutional Review Board of Sapporo Medical University Hospital approved this study (No. 342–201, approved on 02-09-2023).

### IIP biorepository enrollment procedures

Patients enrolled in this ILD registry underwent standardized collection of physical and laboratory evaluation, pulmonary function testing (PFT) and high-resolution computed tomography (HRCT) at the time of diagnosis and enrollment. Each HRCT was interpreted by two pulmonologists specializing in ILD to determine the predominant HRCT pattern. Patients underwent bronchoalveolar lavage (BAL) within three months of enrollment. All patients included in this subsequent analysis were treatment naïve at the time of BALF sample collection. Survival status was extracted from the medical record for each patient and updated at time of data collection by the parent investigators as of November 2023.

BALF was collected following a standardized protocol with 50mL of 0.9% sterile saline being instilled into the right middle lobe or lingula via a wedged bronchoscope. Lavage fluid was then collected via gentle suction and repeated for a total of three lavages (150mL of 0.9% in total). The collected BALF supernatant was combined, centrifuged to remove cells, and frozen at - 80°C for future study. Each participant signed informed consent for serum and BALF samples to be stored and used for future research.

PFTs at baseline (within 3 months of BALF) were extracted from the medical record as measure of baseline disease severity including forced vital capacity % predicted (FVC%) and diffusing capacity for carbon monoxide % predicted (DLCO%).

### Control subjects

Control subjects without lung disease (n=5) had BALF collected from the University of Kansas Asthma and Airway Translational Research Unit biobank. These samples were collected previously from prospective studies where subjects had research-protocol PFTs and HRCT confirming the absence of ILD and extensive collection of clinical and demographic variables. All subjects underwent informed consent for the parent studies which included consent for banking samples for future research. BALF was obtained using similar methods as discussed above.

### Protein isolation and quantification

BALF samples were thawed and centrifuged at 5000 x g for 5 minutes. Pierce BCA protein assay (ThermoFisher Scientific) was used to quantify the concentration of proteins present in BALF following the manufacturer’s protocol against a standard curve of BSA from 25 – 2000 µg/mL.

A 100 µL aliquot of BALF was reduced with TCEP (5 mM) and incubated at 37°C for 30 minutes. Reduced samples were alkylated with iodoacetamide (10 mM) and incubated in the dark at room temperature for 30 minutes. Ice cold acetonitrile (ACN) was added to each sample to a volume ratio of 3:1. Samples were incubated at -20°C overnight and were subsequently centrifuged at 14,000 x *g* at 4°C for 30 minutes to pellet the proteins. The supernatant was removed, and the pellet was air dried on benchtop for 10 minutes. The proteins were resuspended in 50 mM TEAB pH 8 and digested with trypsin (500 ng) overnight at 37°C with shaking at 500 RPM (Thermomixer, Eppendorf). The digestion was quenched with the addition of formic acid to a final concentration of 1%. Digested samples were stored at -20°C until mass spectrometry analysis. Peptide concentration was measured using a Nanodrop spectrophotometer (Thermo Scientific) at 205 nm prior to LC-MS/MS analysis.

Samples were injected using the Vanquish Neo (Thermo) nano-UPLC onto a C18 trap column (PepMap ™ Neo, 0.3 x 5 mm, 5 µm C18) prior to elution onto the separation column (PepMap™ Neo, 2 µm, 75 µm x 150 mm). Peptides were eluted with a linear gradient (Supplemental Table 1) from the nano-LC directly interfaced with the Orbitrap Ascend mass spectrometer (Thermo) equipped with a high field asymmetric waveform ion mobility spectrometry (FAIMS) source using 3 compensation voltages. Details can be found in the supplemental material. Raw files were searched against the human protein database downloaded from Uniprot on 05-05-2023 using SEQUEST in Proteome Discoverer 3.0 [11]. During the search, peptide groups and protein abundances were normalized by applying the normalization of the total abundance values for each channel across all files, equalizing the total abundance between different runs. Details can be found in supplemental materials. Results were exported to Microsoft Excel for further analysis in R.

Results were exported to Microsoft Excel for further analysis in R.

### Statistical Analysis

To delineate lung-based molecular phenotypes and assess their clinical relevance, we first clustered subjects IIP subjects (n = 23) based on their protein expression signature. This was carried out using *k*-means clustering based on the log-transformed expression values of only those proteins that were detected in all 23 IIP subjects. The optimal number of clusters (*k*) was determined by applying the NbClust methodology [12], as implemented in the NbClust R package, to the matrix of log-transformed protein expression data, using only the proteins that were detected in all 23 IIP subjects. The NbClust framework and corresponding R package provide 30 different clustering indices (e.g., silhouette) to assist in the determination of the optimal number of clusters when unsupervised clustering is applied to a data set. The resulting clusters were next examined to determine their association with various clinical and epidemiological variables, including: age, gender, smoking pack years, FVC%, DLCO %, clinical diagnosis (IPF vs. IPAF) and radiologic ILD pattern (i.e., NSIP, OP, PPFE, UIP), and GAP index [13, 14]. The association between cluster membership and continuous variables (age, smoking pack years, FVC%, and DLCO%) and categorical variables (radiologic patterns, clinical diagnosis and GAP index) were assessed using a series of Kruskal-Wallis and Fisher’s exact tests, respectively. We also determined the specific proteins that uniquely discriminated the identified *k*-means clusters by conducting formal differential protein expression analyses between each pair of the identified clusters. This was carried out by fitting a series of linear regression models, modeling log-transformed expression as the dependent variable against cluster membership as the independent variable, and adjusted for subject age, gender, smoking pack years, FVC%, and subsequent treatment. Models were fit independently to each of the 302 proteins that were observed/detected across all 23 IIP subjects. Proteins that uniquely and uniformly discriminated the identified clusters were subjected to an overrepresentation analysis (ORA) to determine biological pathways and GO terms that are significantly overrepresented with such proteins. The latter was carried out using the enrichGO function in the clusterProfiler Bioconductor package. To assess the potential clinical relevance of the identified *k*-means clusters, we next examined the association between cluster membership and overall survival using a multivariable Cox proportional hazards model adjusted for age, smoking pack years, FVC%, and clinical diagnosis. Time to death/censoring was calculated as the amount of time elapsed from BALF collection to death or censoring.

## Results

The IIP cohort mean age was 70 (63-75) years and predominantly male (60.9%) with median cigarette smoking history of 27 pack-years (0.25-48). 5 patients in the IIP cohort had IPF, while 18 met IPAF criteria. The median baseline FVC% in the IIP group was 84.5 (74-94.1) and median DLCO was 52.4 (45.3-60.4) which were both lower than the control group (Table 1).

**Table 1.** Patient characteristics.

| Variable | IIP (n=23) | Controls (n=5) | p-value |
| --- | --- | --- | --- |
| Age, median (IQR)) | 70 (63-75) | 67 (62-72) | 0.88 |
| Male, n (%) | 14 (60.9%) | 3 (60%) | 0.65 |
| Pack Years, median (IQR) | 27 (0.25-48) | 35 (0.45-45) | 0.59 |
| FVC%, median (IQR) | 85.4 (74-94.1) | 109.2 (105.1-120.3) | 0.09 |
| DLCO%, median (IQR) | 52.4 (45.3-60.4) | 65 (54.6-76.3) | 0.28 |
| GAP index score, mean (SD) | 2.9 (0.92) | na |  |
| Vital status, Alive, n (%) | 12 (52.2%) |  |  |
| Time to death, years (median) (IQR) | 3.83 (1.81-29.86) | 3.22 (3.17-3.67) | 0.70 |

IIP: idiopathic interstitial pneumonias, FVC%: forced vital capacity percent predicted, DLCO%: diffusion capacity for carbon monoxide percent predicted, GAP: gender, age, physiology index for survival, IQR = interquartile range

Over a median follow-up of 6.23 (1.81-29.86) years 11 deaths occurred.

### Overall Heatmap

To verify that this method could differentiate IIP samples from control samples, we compared BALF protein expression across 23 IIP patients and five healthy control samples. There were 1176 proteins detected at least once in the samples (Supplemental Table 2). Among these, 45 proteins were considered contaminants and excluded from analysis. A heatmap of a subset of 50 proteins that were most significantly, differentially expressed (lowest p-value) demonstrated the different protein abundance profiles by disease or control (Figure 1-A). Further, principal component analysis visibly discriminated between control and IIP samples (Figure 1-B), with the first two principal components accounting for 38.9% of the variance, indicating a distinct protein expression profile associated with the disease state.

**Figure 1:**
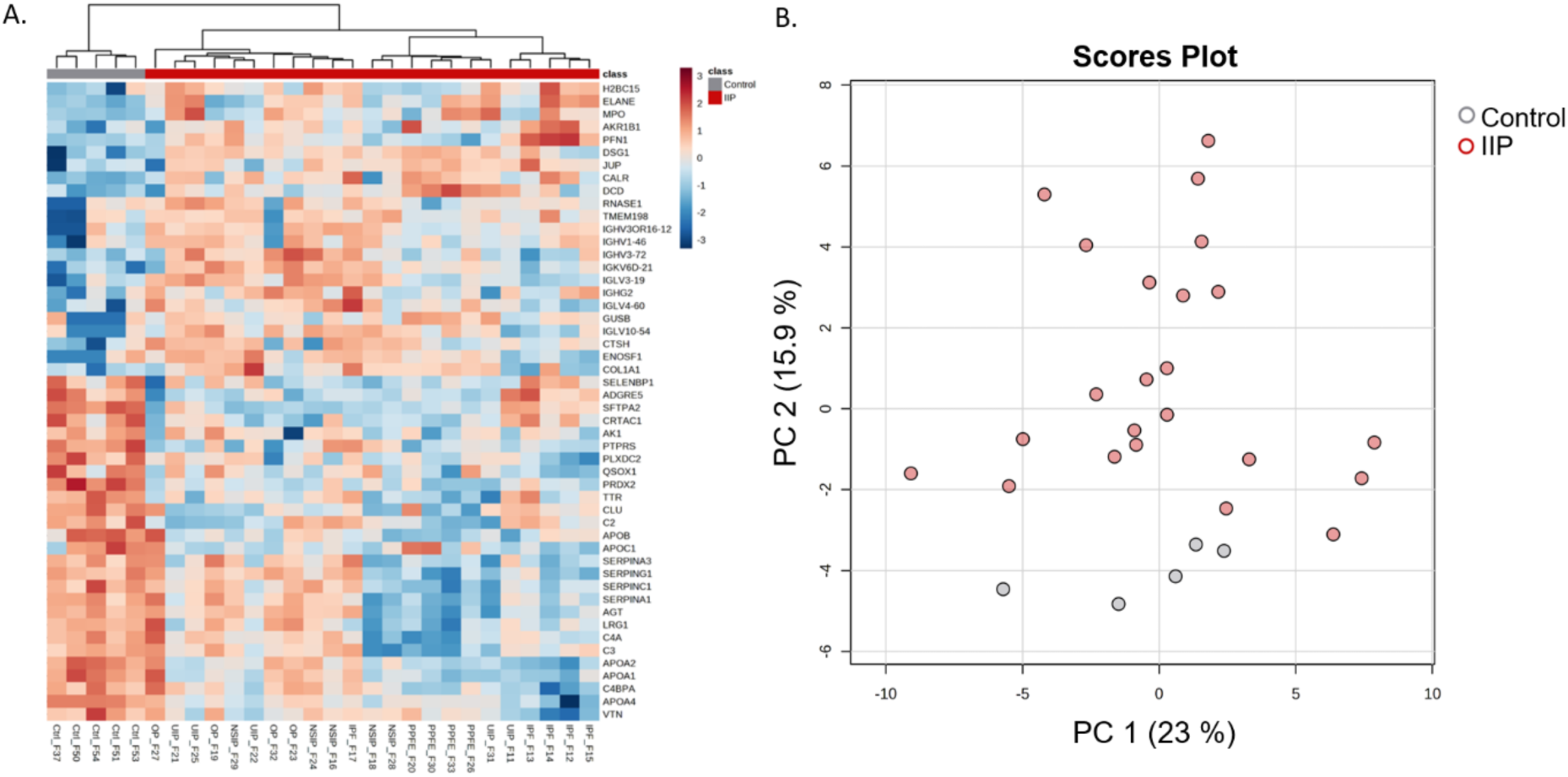
A. An abbreviated heatmap of protein expression across controls and idiopathic interstitial pneumonia (IIP) patients. The top fifty differentially expressed proteins defined by having at least two times fold change (FC) in either direction and p-value <0.05 for t-test between IIP and control samples are shown in Figure 1A. Gray bars on the top row indicate control samples while red bars indicate IIP patients. The dendrogram above the heatmap indicates two primary distinct branches (controls vs IIP). B. Principal component analysis (PCA) plot with the same color coding for each patient bronchoalveolar lavage fluid (BALF) protein profile (grey = controls and red = IIP).

### IIP Cluster Analysis

Our cluster analysis was restricted to only those proteins that were detected in all 23 IIP subjects resulting in a total of 302 proteins in the *k*-means clustering analysis. The appropriate number of possible unique protein expression clusters across the IIP group as determined by the NbClust function [12] in R was three. A heatmap of the protein abundances by cluster is shown in Figure 2. These unsupervised protein cluster memberships [Cluster 1 (n = 8), Cluster 2 (n = 5), and Cluster 3 (n = 10)] were then subjected to a series of Kruskal-Wallis tests to examine the association between cluster membership and selected demographic and clinical characteristics, none of which were statistically significant (smoking pack years: p-value = 0.86, age: p-value =0.71, FVC% predicted: p-value = 0.068, and DLCO % predicted: p-value = 0.44) suggesting that these clinical or demographic factors do not primarily drive the clustering of patients in this study (Supplemental Table 3 and Supplemental Figure 1).

**Figure 2:**
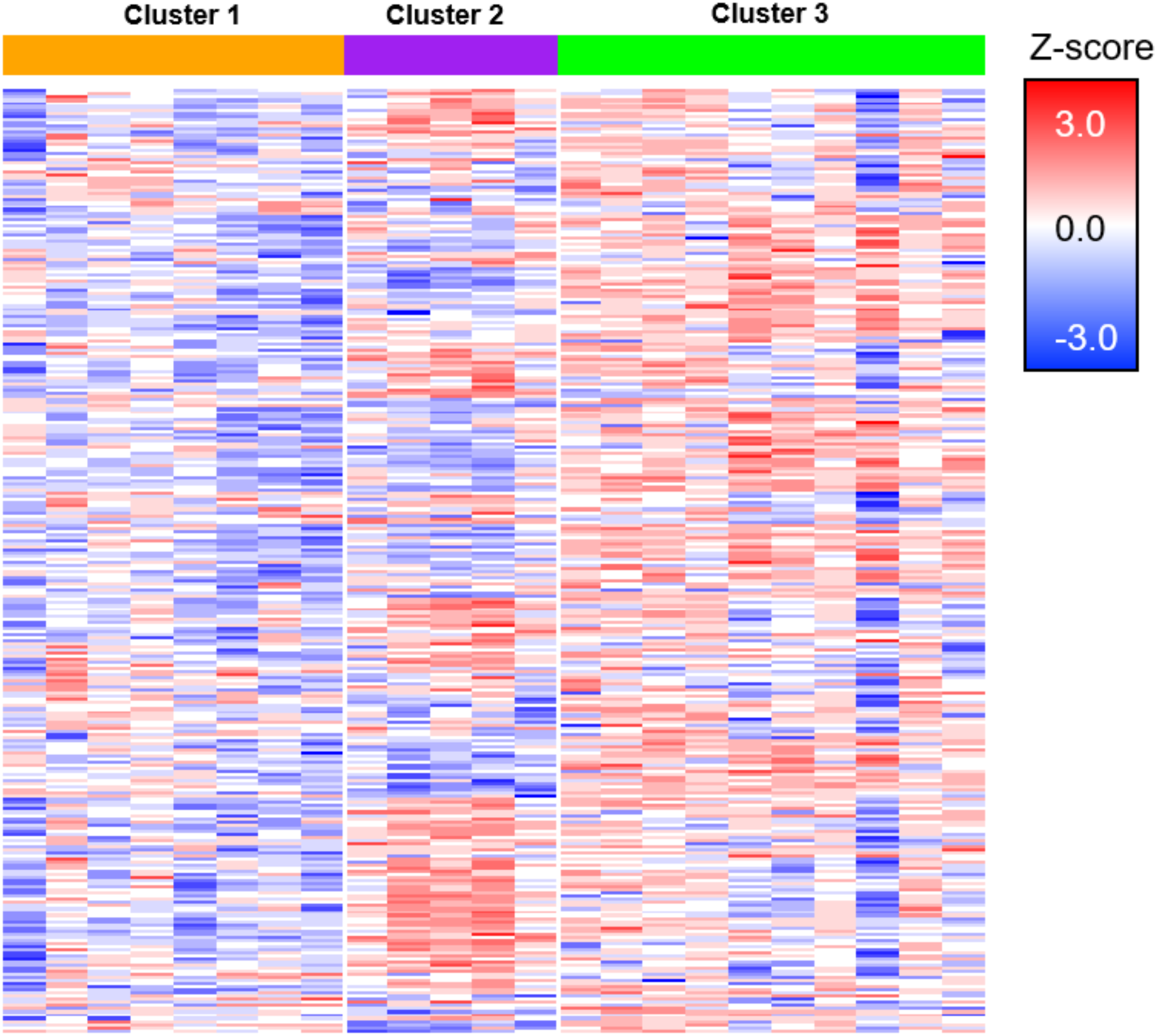
Protein expression heatmap of the 302 proteins across the 23 IIP patients. *k*- means cluster analysis and majority rule using NbClust function in R determined 3 as the appropriate number of clusters. In this heatmap the patients are represented in columns and color-coded for cluster designation on the top row. Cluster 1 in orange, Cluster 2 in purple and Cluster 3 in green.

### Association between cluster membership and survival

Compared to Cluster 1, which had the worst survival, patients in Clusters 2 and 3 were observed to have a reduced hazard of death in models adjusting for age, smoking pack-years, and FVC%, HR = 0.201, CI: (0.039, 1.044) and HR = 0.092, CI: (0.017, 0.479), respectively (Figure 3). This association between survival and cluster designation remained significant when adding subsequent immunosuppression treatment to the model (with pack-years, gender, age, and FVC% (p-value = 0.032).

**Figure 3:**
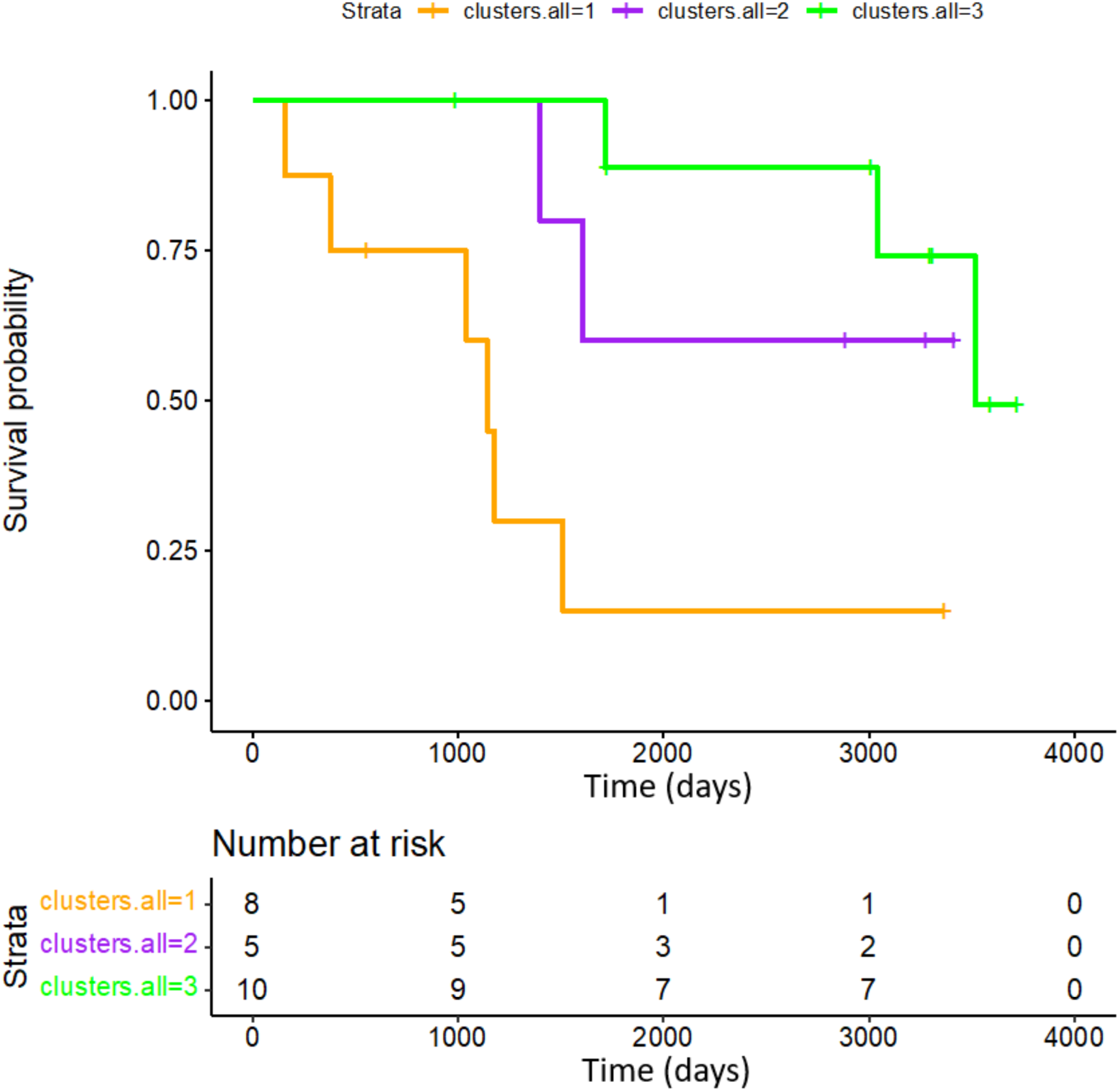
Cox-proportional hazards analysis for each cluster and time to survival in days plotted on x-axis and survival probability plotted on y-axis. Cluster 1 is orange, Cluster 2 is purple, and Cluster 3 is green.

To assess the impact of these novel, unsupervised BALF protein clusters on survival prediction we compared the concordance-index (C-index) of cluster membership with different combinations of risk factors. In this analysis, cluster membership alone (C-index: 0.79) outperformed models with age (C-index: 0.71); the GAP index (C-index: 0.71). When cluster membership and the GAP index were combined in a single model, the C-index increased to 0.88, indicating increased prognostic performance compared to models that include only cluster membership or GAP index (Figure 4).

**Figure 4:**
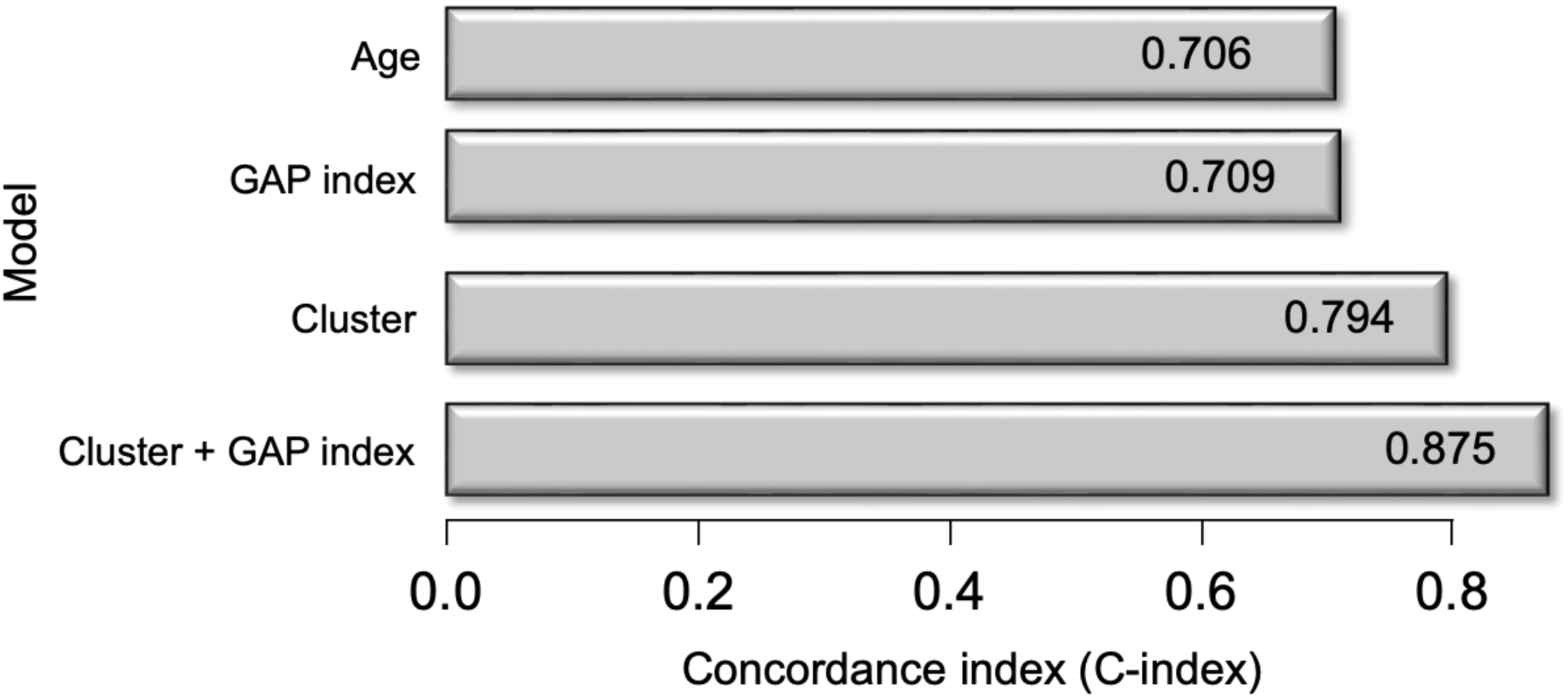
Protein cluster (Cluster) identity outperforms GAP index and age for C-index for survival. When cluster membership and the GAP index were combined in a single model, the C-index increased to 0.88.

### IPF and IPAF radiographic pattern within each cluster

Cluster 1 (worst survival group) included: two patients with IPAF-NSIP, all four IPAF-PPFE patients and two patients with IPAF-UIP. Cluster 2 (intermediate survival group) included four patients with IPF, and one patient with IPAF-UIP. Cluster 3 (best survival group) included all four IPAF-OP patients, three patients with IPAF-NSIP, two patients with IPF-UIP and one patient with IPF (Figure 5) (Supplemental Table 4 for clinical and demographic characteristics based on cluster designation).

**Figure 5:**
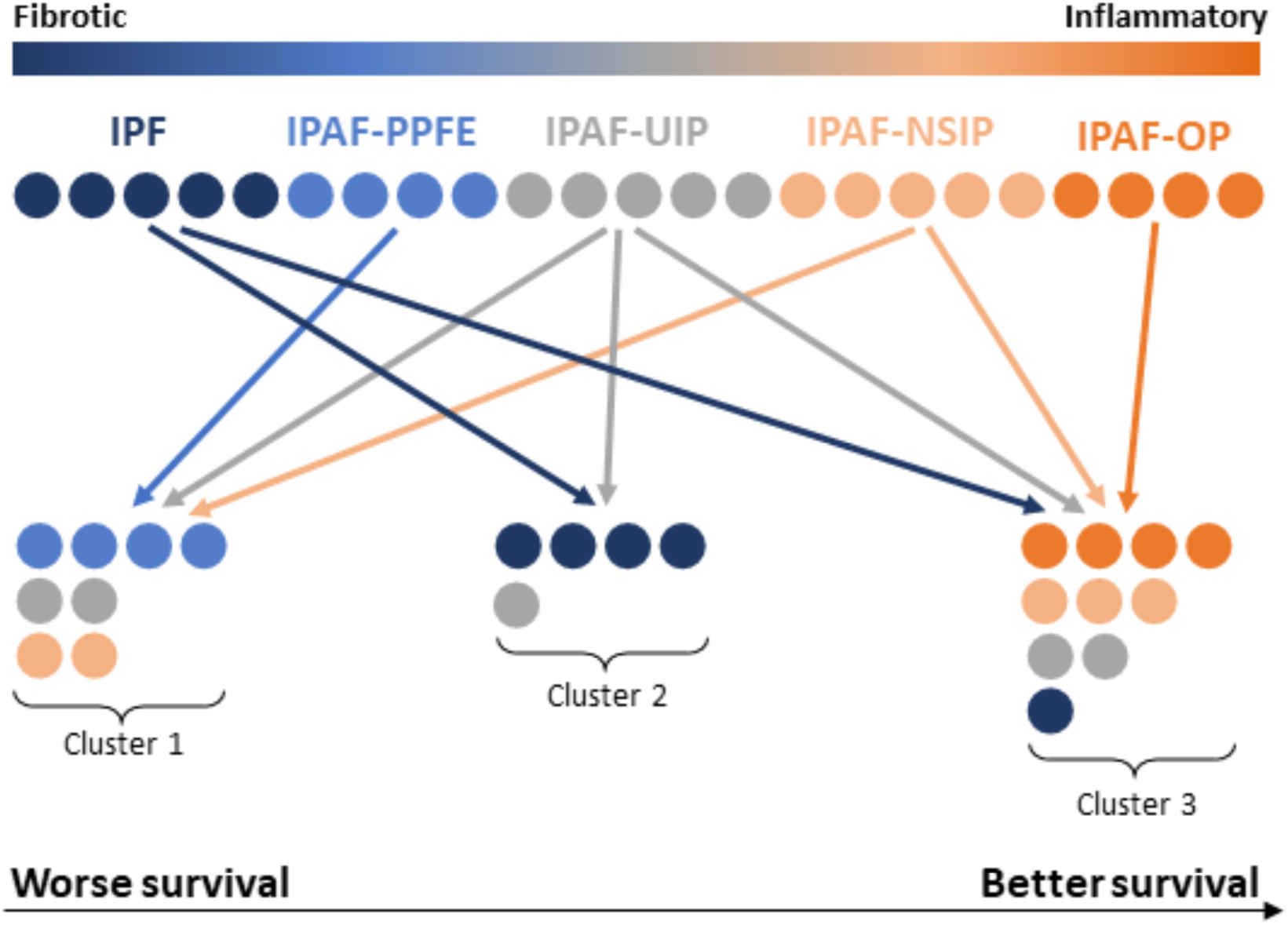
Patient diagnoses of idiopathic interstitial pneumonia (IIP) based on radiographic pattern, thought to be more fibrotic (idiopathic pulmonary fibrosis (IPF), pleuorparenchymal fibroelastosis (IPAF-PPFE), and usual interstitial pneumonia (IPAF-UIP)) or more inflammatory (non-specific interstitial pneumonia (IPAF-NSIP) and organizing pneumonia (IPAF-OP)). The dispersion of each patient in the analysis from IIP radiographic designations based on the unsupervised bronchoalveolar lavage (BALF) proteomics clustering data is shown by arrows and similar color scheme.

### Uniquely discriminating protein analysis of Cluster 1 versus Clusters 2 and 3

Given that Cluster 1 was associated with the worst survival trajectory, we performed a discrimination analysis to determine proteins that uniquely discriminated Cluster 1 from Clusters 2 and 3. In this analysis (Figure 6), there was a notable linear relationship seen across the top 10 discriminating proteins whose expression levels were decreased in Cluster 1 (worst survival) compared to Cluster 3 (best survival). These 10 proteins are similarly involved in inflammatory signaling via corticosteroid receptor activity (SERPINA6, SERPINA1) and complement system activation (C3, F2, SERPIND1, SERPINC1, C5, C9, C8B, C8A) [15, 16].

**Figure 6:**
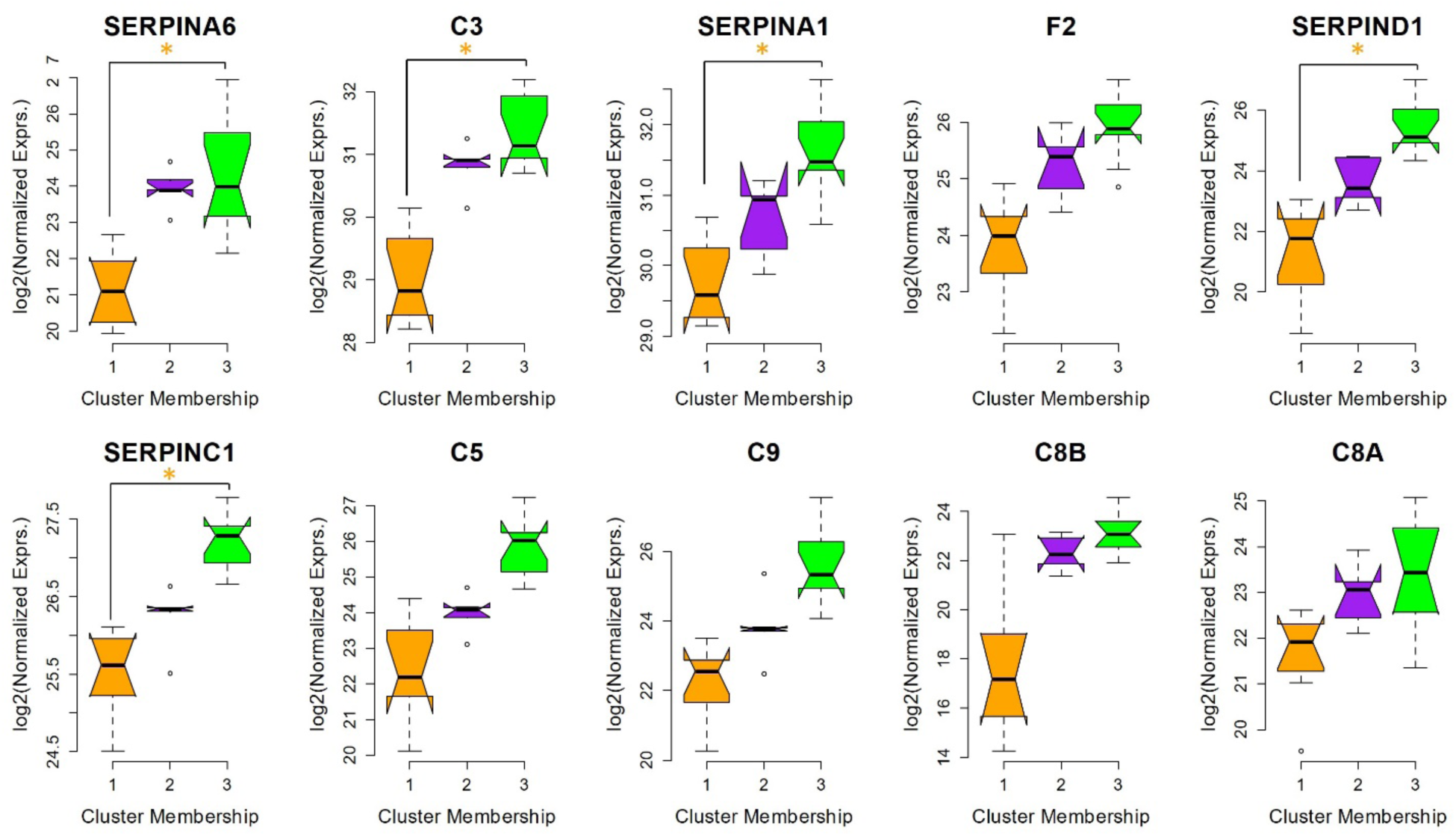
Notched box r plots of the top 10 discriminating proteins that increase in expression from worst survival cluster (Cluster 1, orange), intermediate survival (Cluster 2, purple), and best survival (Cluster 3, green). The median normalized abundance is shown by the thick black line with the minimum and maximum indicated by the whiskers. The top and bottom lines of the box indicate 75^th^ percentile and 25^th^ percentile, respectively. These 10 proteins are similarly involved in inflammatory signaling via corticosteroid receptor activity (SERPINA6, SERPINA1) and complement system activation (C3, F2, SERPIND1, SERPINC1, C5, C9, C8B, C8A) [15, 16]. *p-value < 0.05

In survival analyses adjusted for age, gender, smoking pack years, baseline FVC% and IPF diagnosis, the abundance of each of these proteins was found to a hazard ratio for survival < 1, suggesting an inverse relationship between their expression and hazard of death (Supplemental Table 5). Of these 10 proteins, five had significant associations with survival after adjusting for those factors (Table 2).

**Table 2.** Five uniquely discriminating proteins (Cluster 1 vs. Clusters 2 and 3) with significant association with survival after adjustment for age, sex, smoking status and baseline severity (FVC%).

| Protein Name | HR | CI | p-value |
| --- | --- | --- | --- |
| **SERPINA6** | 0.41 | (0.21, 0.82) | 0.012 |
| **C3** | 0.34 | (0.13, 0.91) | 0.031 |
| **SERPINA1** | 0.37 | (0.14,0.98) | 0.045 |
| **SERPIND1** | 0.67 | (0.46, 0.97) | 0.032 |
| **SERPINC1** | 0.38 | (0.16, 0.95) | 0.039 |

### Uniformly discriminating proteins

A total of 19 proteins were observed that uniformly discriminated all three clusters, i.e., proteins that exhibited most significant differential expression across all three clusters (Figure 7 and Supplemental Figure 2). We incorporated these 19 proteins in a pathway analysis using an over-representation analysis (ORA). Using these 19 uniformly discriminating proteins across all three clusters, the ORA (Figure 8) revealed a significant enrichment of pathways involved in the immune response, particularly those associated with immunoglobulin production, B cell-mediated immunity, and lymphocyte-mediated immunity. Pathways related to the production of molecular mediators of the immune response were also overrepresented. Notably, adaptive immune responses, including somatic recombination of immune receptors built from immunoglobulin superfamily domains, were prominent, suggesting active engagement of specific and adaptive defense mechanisms (p-values < 0.004 for all pathways mentioned).

**Figure 7:**
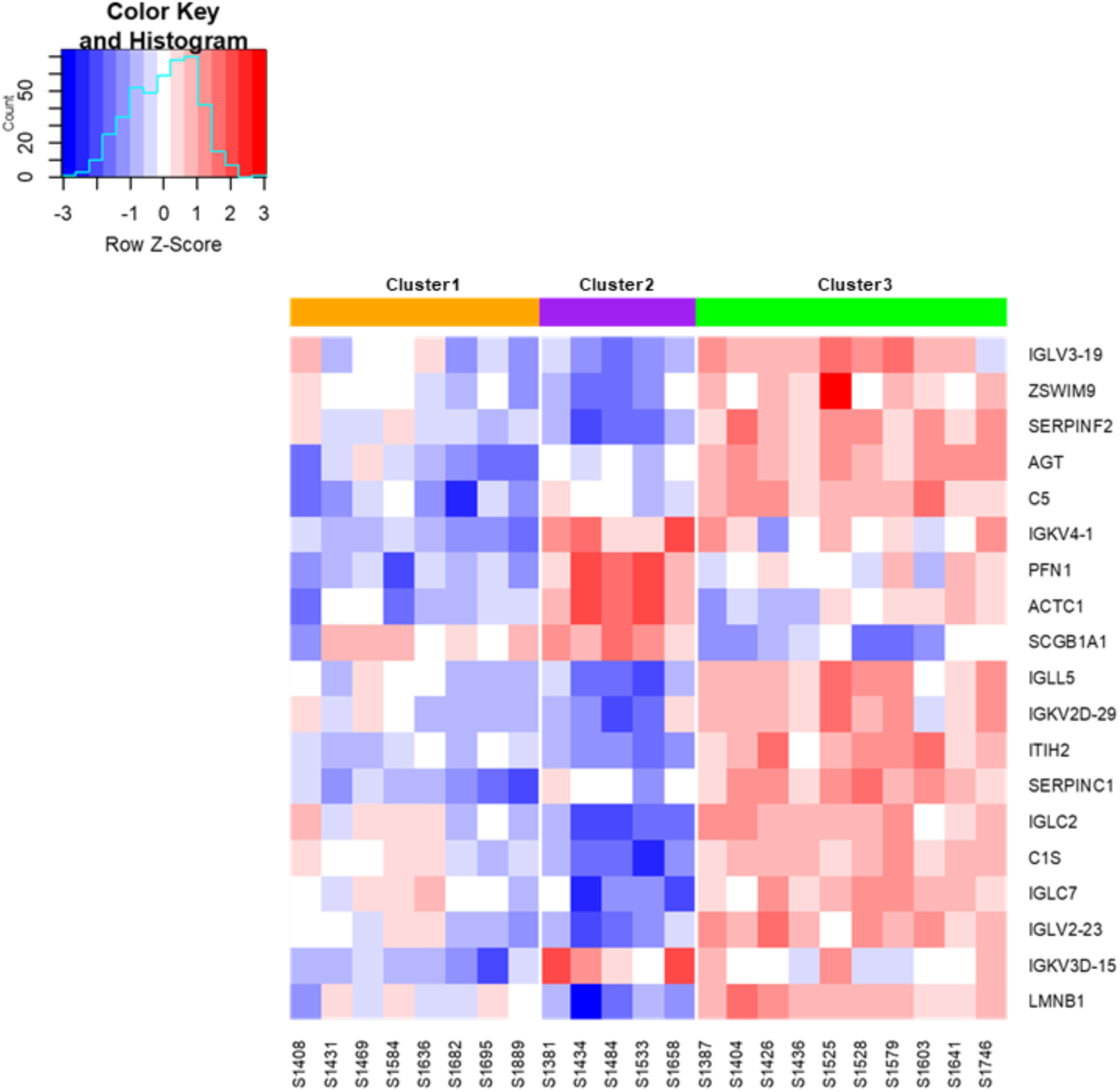
Heatmap of proteins uniformly discriminating across three clusters.

**Figure 8:**
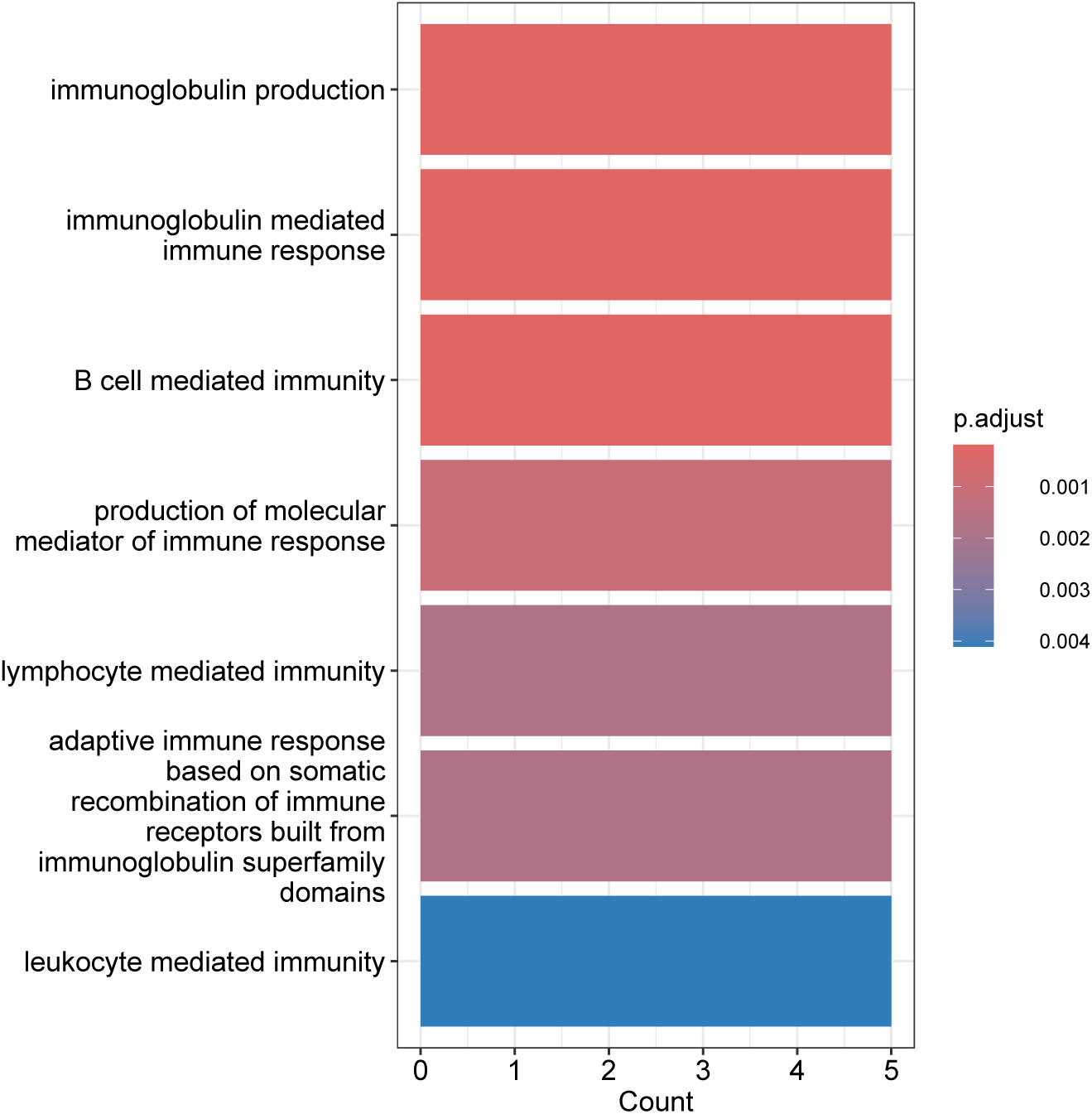
Overrepresentation analysis of proteins uniformly discriminating clusters 1, 2 and 3.

## Discussion

Proteomics is a high-throughput, systems biology approach that can identify and quantify proteins within a given sample, thus providing a comprehensive overview of the cellular and molecular mechanisms at play. In the context of lung disease, proteomics enables the characterization of protein expression profiles that may reflect the underlying pathophysiology of heterogenous conditions such as ILD. In this study, we isolated a cohort of subjects with IPF and IPAF across a diverse array of radiographic patterns with banked bronchoalveolar lavage fluid (BALF) that was collected before treatment initiation to perform label-free, quantitative proteomics of the lung microenvironment.

BALF reflects the protein signatures of the diseased lung parenchyma more accurately than serum or plasma and offers a broader representation of lung phenotypes than isolated tissue samples which are subject to heterogeneity and sampling error. BALF has been shown to be the most direct measure of the lung microenvironment as compared to other body fluids, such as serum, sputum, and nasal lavage fluid [17–19]. BALF has an advantage over surgical lung biopsy due to its inherent safety [20], where mortality after surgical lung biopsy approaches 2% in US populations [21]. Moreover, despite the technical challenges associated with bronchoscopy, BALF remains accessible for ongoing clinical research, facilitating the prospective study and validation of molecular phenotypes within the lung microenvironment.

Previous studies have explored the BALF proteome in individual ILD disease states, largely focused on IPF [22–29], while our analysis was focused on identifying novel proteomics clusters across a group of patients with IIP.

Proteins are a major component of BALF, and those specific to pulmonary pathology such as mucins and surfactant proteins have been found in high amounts in BALF [30, 31]. BALF are known to contain endogenous proteins and peptides that exhibit biological activity specific in the distal lung [32].

In this study, we highlighted a panoramic strategy to leverage the powerful systems biology approach of proteomics to identify novel molecular profiles across a spectrum of IIP, thus enabling us to identify clusters that are driven by lung microenvironment biology as opposed to visual radiographic subtypes and subjective diagnostic criteria.

Despite the small sample size, these unsupervised pretreatment BALF protein profile clusters largely recapitulated decades of clinical experiences. For instance, all four patients with IPAF and organizing pneumonia clustered together in the “inflammatory” cluster (Cluster 3) with best survival. Similarly, four out of five IPF subjects grouped together in Cluster 2 and all four patients with IPAF-PPFE were included in the cluster with the worse survival (Cluster 1).

The current state of the art for prognostication in ILD such as IPF or IPAF remains a clinical model that combines age, gender and two physiologic variables to create a risk score for mortality that has been validated in many forms of ILD (GAP index) [13, 14]. In these current data, unsupervised BALF protein cluster designation outperformed the GAP index and further improved prognostic accuracy when combined with GAP index. This indicates a potential prognostic role for this approach that calls for further validation in larger datasets and across other ILD types.

The possible clinical utility of this approach is intriguing not only for prognostication, but there are also exciting translational implications for drug discovery and repurposing. For instance, proteins that uniquely discriminated clusters exhibited an intriguing linear relationship between inflammatory signaling and complement activation, mimicking a protective dose-response for survival. The possibility of a linear, continuous marker in BALF, agnostic to radiographic or etiology that is associated with survival offers the first glimpse for the potential of this method to guide precision-medicine, patient specific interventional trials.

The potential implications of this approach, if applied broadly, are perhaps most evident when considering the five patients with IPAF-UIP in this analysis. These five IPAF-UIP patients were separated across all three clusters, which led to the hypothesis that while some underlying etiologies and radiographic patterns might have very similar BALF protein expression patterns (IPAF-PPFE and IPAF-OP), there are some patients with visually indiscernible differences in lung phenotypes that drive their clinical outcomes. The current approach to making decisions on diagnosis and treatment for individual patients with IPAF-UIP is driven largely by practice patterns and unvalidated patient characteristics. There has yet to be a clinical marker that can reliably predict treatment response or survival for these patients. These data highlighted the potential of this approach if expanded and validated across many forms of ILD to identify unique endotypes of ILD for targeted treatment.

There are important limitations to these data. Because of the small sample size, the observed associations between survival trajectory and protein expression clusters remain hypothesis generating until validated in larger, more diverse datasets. It is important to consider how this unsupervised cluster analysis would evolve with increased sample size and with a larger breadth of diagnoses, such as established autoimmune ILD or hypersensitivity pneumonitis. Other considerations that limit broad application of these results relate to the single-center study design with important limitations to generalizability due to the single ethnicity in our IIP cohort from Japan compared to a healthy US control cohort. This lack of patient diversity limits applicability of the findings, and BALF collection methods may vary from site to site, which could limit the external validity of this approach in future multi-site studies.

There are limitations to the utility of this approach for clinical uptake based on the current lack of clinical indications for bronchoscopy in these parenchymal lung diseases. Further work to validate and extend these observations will need to assess if less invasive compartments such as induced sputum, exhaled breath condensate, or blood could recapitulate these findings.

However, one possibility of validation of these findings in larger, multiethnic cohorts would be increased clinical utility of bronchoscopy in these scenarios and future prospective work would need to assess and test the balance of harms and benefits in IIP.

In summary, these current results offer a potential avenue to move diagnostic and treatment choice for individual patients forward through the discovery of lung microenvironment endotypes that drive clinical outcomes. Further studies are required to realize this potential; however, diagnosis and treatment choice for patients with ILD should be guided by authentic, molecular markers of disease activity rather than reliant solely on subjective visual and clinical features.

These results offer a potential path towards that important effort.

## Data Availability

All data available upon reasonable request to authors

