## Supplement for "Proteomic profiling of bronchoalveolar lavage fluid uncovers protein clusters linked to survival in idiopathic forms of interstitial lung disease"

**Supplemental Table 1.** Peptide elution gradient

| Time (min) | Flow Rate | %B |
|---|---|---|
| **5** | 0.35 µL/min | 2 |
| **105** | 0.35 µL/min | 25 |
| **125** | 0.35 µL/min | 40 |
| **140** | 0.35 µL/min | 100 |
| | 0.35 µL/min | 2 |

Peptide elution method. Mobile phase A: 0.1% formic acid (FA) in water; mobile phase B: 80% ACN with 0.1% FA in water.

**Supplemental Method.**

Intact peptides were detected in the Orbitrap at 120,000 resolving power from 375-1500 *m/z*. Peptides with charge +2-7 were selected for fragmentation by higher energy collision dissociation (HCD) at 28% NCE and were detected in the ion trap with rapid scan rate. FAIMS voltages were set at -45 (1.4 s), -60 (1 s), -75 (0.6 s) CV for a total duty cycle time of 3 s. Dynamic exclusion was set to 60s after one instance and mass list was shared between the FAIMS voltages. Source ionization was set at 1700 V with the ion transfer tube temperature at 305 °C.

Technical normalization: Raw files were searched against the human protein database downloaded from Uniprot on 05-05-2023 using SEQUEST in Proteome Discoverer 3.0 [11]. During the search, peptide groups and protein abundances were normalized by applying the normalization of the total abundance values for each channel across all files, equalizing the total abundance between different runs. Detailes can be found in supplemental materials. Specifically, sums the peptide group abundances for each sample and determines the maximum sum for all files. The normalization factor is the factor of the sum of the sample and the maximum sum in all files.

**Supplemental Table 2**. Global protein identification (attached xls file)

**Supplemental Table 3**. Patient demographic and clinical characteristics

| Variable | IPF (n=5) | IPAF-UIP (n=5) | IPAF-PPFE (n=4) | IPAF-NSIP (n=5) | IPAF-OP (n=4) | p-value (ANOVA) |
|---|---|---|---|---|---|---|
| Age, median (IQR) | 67 (62-72) | 74 (70-76)) | 74 (70-76)) | 53 (38-66) | 70 (59.5-77.3) | 0.073 |
| Sex, Male, n (%) | 3 (60%) | 3 (60%) | 3 (75%) | 3 (60%) | 2 (50%) | na |

| | | | | | | |
|---|---|---|---|---|---|---|
| Pack Years, median (IQR) | 35 (0.45-45) | 45 (10-48) | 38.5 (15-57.6) | 20 (18.8-55.5) | 13.5 (0-41.25) | 0.896 |
| FVC% predicted, median (IQR) | 109.2 (105.1-120.3) | 90 (88.3-90.2) | 82.7 (72.63-92.78) | 71.5 (69.2-78.9) | 78.3 (70.9-80.2) | 0.003 |
| DLCO% predicted, median (IQR) | 56 (54.6-76.3) | 47 (45.3-61.8) | 59.9 (52.1-67.1) | 44.7 (42.8-53.8) | 46.1 (45.8-48.6) | 0.397 |
| GAP index score, mean (SD) | 2.8 (0.8) | 3.2 (1.1) | 2.3 (0.6) | 2.8 (1.3) | 3.3 (0.5) | 0.71 |
| Vital status, Alive | 2 (40%) | 1 (20%) | 3 (75%) | 4 (80%) | 2 (50%) | na |
| Time to death, days (IQR) | 1175 (1159-1342.5) | 1503 (1088.8-1633.3) | 1041 | 3039 | 1947 (1162-2732) | 0.574 |
| IPAF criteria<br>Morphology<br>Serology<br>Clinical | <br>0/5 (0%)<br>1/5 (20%)<br>1/5 (20%) | <br>0/5 (0%)<br>5/5 (100%)<br>5/5 (100%) | <br>4/4 (100%)<br>4/4 (100%)<br>0/4 (0%) | <br>5/5 (100%)<br>5/5 (100%)<br>2/5 (40%) | <br>4/4 (100%)<br>2/4 (50%)<br>2/4 (50%) | na |

IPF: idiopathic pulmonary fibrosis, IPAF: interstitial pneumonia with autoimmune features, UIP: usual interstitial pneumonia, PPFE: pleuroparenchymal fibroelastosis, NSIP: non-specific interstitial pneumonia, OP: organizing pneumonia, FVC: forced vital capacity, DLCO: diffusion capacity for carbon monoxide, GAP: gender, age, physiology index for survival.

Supplemental Table 4. Patient and clinical characteristics by clusters:

| Clinical variables | Cluster 1 (n=8) | Cluster 2 (n=5) | Cluster 3 (n=10) | p-value (ANOVA unless marked by*) |
|---|---|---|---|---|
| Age, median (IQR) | 71 (65.5-70) | 65 (62-72) | 69.5 (55.8-75.5) | 0.919 |
| Sex, male, n (%) | 6 (75%) | 3 (60%) | 5 (50%) | 0.9747 |
| Pack years, median (IQR) | 19.4 (7.5-55.9) | 45 (0.45-45) | 31 (5-75) | 0.563 |
| FVC% predicted, median (IQR) | 90.1 (72.6-92.8) | 105.1 (88.3-12.3) | 78.3 (71.5-82.1) | 0.0171 |
| DLCO% predicted, median (IQR) | 55.8 (44.9-62.5) | 56 (54.6-76.3) | 46.3 (45.4-51) | 0.432 |
| GAP index score, mean (SD) | 3.1 (1) | 2.2 (0.8) | 3.1 (0.8) | 0.257 |
| Vital status, Alive, n (%) | 2 (25%) | 3 (60%) | 7 (88%) | 0.8976 |
| Time to death, days, median (IQR or range*) | 1092 (543-1167) | 1503 (1400-1606)* | 3039 (1715-3517)* | 0.00941 |
| Time to death, years, median (IQR or range*) | 3 (1.5-3.2) | 4.1 (3.8-4.4)* | 8.3 (4.7-9.6)* | 0.00941 |
| IPAF criteria, n (%)<br>Morphology<br>Serology<br>Clinical | <br>6/8 (75%)<br>8/8 (100%)<br>3/8 (38%) | <br>0/5 (0%)<br>2/5 (40%)<br>2/5 (40%) | <br>7/10 (70%)<br>7/10 (70%)<br>5/10 (50%) | N/A |
| Underwent subsequent immunosuppressive treatment | 1/8 (12.5%) | 1/5 (20%) | 6/10 (60%) | 0.1215* |

*Fisher's Exact Test

**Supplemental Figure 1**. Kruskal-Wallis tests

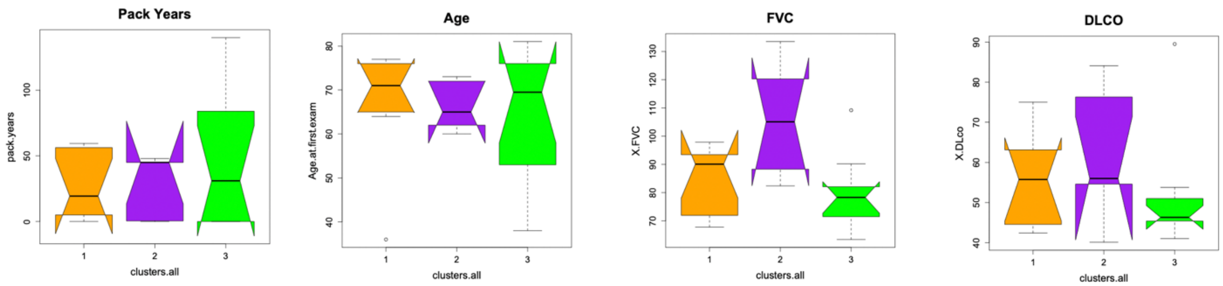


p-value = 0.8633 p-value = 0.7098 p-value = 0.0680 p-value = 0.4444

Supplemental Figure 1 Legend. Violin plots of Kruskal-Wallis tests examining the association between cluster membership and selected demographic and clinical characteristics (smoking pack years, age, FVC%y, DLCO%). Colors represented Clusters (Cluster 1 – orange, Cluster 2 – purple, Cluster 3 – green).

**Supplemental Table 5**. Hazard ratios of proteins uniquely discriminate Cluster 1 from Clusters 2 and 3.

| | HR | 95%CI | pval |
|---|---|---|---|
| SERPINA6 | 0.4101034 | (0.2048, 0.8213) | 0.01188177 |
| C3 | 0.3387911 | (0.1267, 0.9059) | 0.03101418 |
| SERPINA1 | 0.3698145 | (0.1401, 0.9762) | 0.04457449 |
| F2 | 0.5862551 | (0.2995, 1.1477) | 0.11921609 |
| SERPIND1 | 0.6665833 | (0.4602, 0.9656) | 0.03194525 |
| SERPINC1 | 0.3808444 | (0.1526, 0.9508) | 0.03862855 |
| C5 | 0.6289068 | (0.3737, 1.0585) | 0.08080829 |
| C9 | 0.6569248 | (0.4075, 1.0589) | 0.08453386 |
| C8B | 0.8286892 | (0.6312, 1.0879) | 0.17603781 |
| C8A | 0.5676409 | (0.26, 1.2392) | 0.15515416 |

**Supplemental Figure 2**. Uniformly discriminating proteins across three clusters.

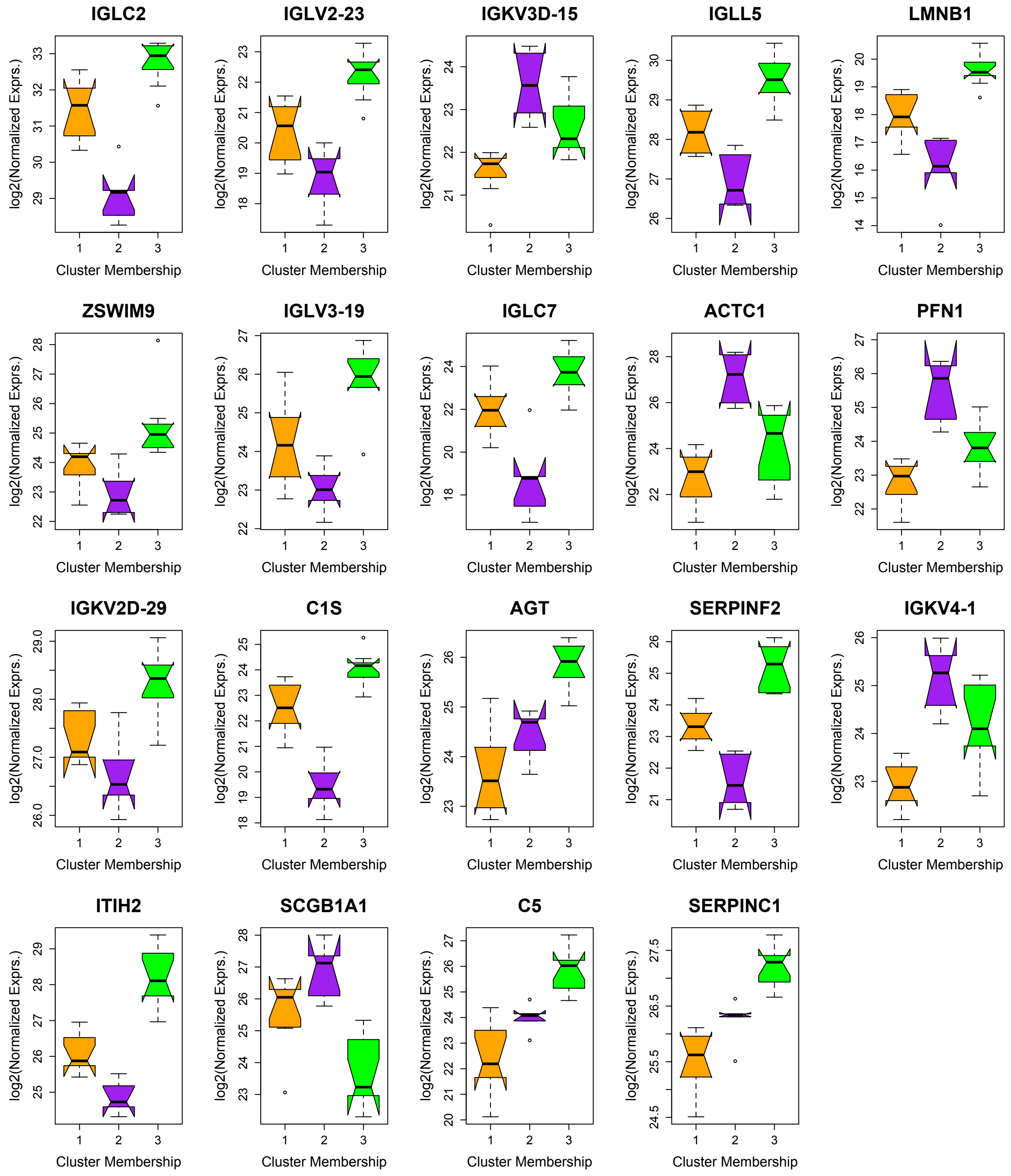


Supplemental Figure 2 Legend. Violin and whisker plot of the 19 uniformly discriminating proteins that had significant discrepancies in expression among Cluster1 (orange), Cluster 2 (purple), and Cluster 3 (green). The mean normalized abundance is shown by the thick black line with the minimum and maximum indicated by the whisker.
